# Application of under-oil open microfluidic systems for rapid phenotypic antimicrobial susceptibility testing

**DOI:** 10.1101/2022.06.24.22276877

**Authors:** Chao Li, Sue McCrone, Jay W. Warrick, David R. Andes, Zachary Hite, Cecilia F. Volk, Warren E. Rose, David J. Beebe

## Abstract

Antimicrobial susceptibility testing (AST) remains the cornerstone of effective antimicrobial selection and optimization in patients. Despite recent advances in rapid pathogen identification and resistance marker detection with molecular diagnostics, phenotypic AST methods remain relatively unchanged over the last few decades. Guided by the principles of microfluidics, we describe the application of a multi-liquid-phase microfluidic system, named under-oil open microfluidic systems (UOMS) to achieve a rapid phenotypic AST. UOMS provides a next-generation solution for AST (UOMS-AST) by implementing and recording a pathogen antimicrobial activity in micro-volume testing units under an oil overlay with label-free, single-cell resolution optical access. UOMS-AST can accurately and rapidly determine antimicrobial activity from nominal sample/bacterial cells in a system aligned with clinical laboratory standards. Further, we combine UOMS-AST with cloud lab data analytic techniques for real-time image analysis and report generation to provide a rapid (i.e., <4 h) sample-to-answer turnaround time, shedding light on its utility as a next-generation phenotypic AST platform for clinical application.

---

Bacterial infection continues to be a major global health threat that has been exacerbated by the emergence and spread of antibiotic-resistant species^1^. The need for rapid (less than a single work shift, i.e., <8 h), reliable AST is crucial for reducing the empiric use of broad-spectrum antimicrobials while ensuring that patients receive timely and adequate treatment^2,3^. The development and use of molecular methods, e.g., quantitative polymerase chain reaction (qPCR)^4,5^, matrix-assisted laser desorption/ionization-time of flight (MALDI-TOF) mass spectrometry (MS)^6^, and Raman spectroscopy^7^, in clinical microbiology laboratories have revolutionized the speed of bacterial identification and antimicrobial resistance detection. However, molecular methods are useful when antimicrobial resistance genes or molecules are known, are currently limited to a few select pathogens and antimicrobials, and do not always predict phenotypic resistance^8^.

Phenotypic methods that directly and functionally screen for bacterial resistance by observing and quantifying the growth or inhibition event of bacteria after exposure to antibiotics provide clinically relevant results and thus remain the gold standard used in clinical settings^8^ (Table 1). The traditional (i.e., manual) standard phenotypic AST methods (including disc diffusion and broth microdilution) require intensive labor and a long detection turnaround time of 1 or 2 days following bacterial identification. Several automated phenotypic AST systems have been approved by the U.S. Food and Drug Administration (FDA) and commercialized. Compared to the manual standard ASTs, the automated systems use integrated software to interpret the AST results and allow real-time report generation and integration into the electronic medical record. These automated systems significantly reduce labor and time (3.5-16 h) of data collection and quantification of antibiotic susceptibility, and today these are used in most large clinical microbiology laboratories. While improving upon the historical standards to provide more rapid AST results, reporting errors and discrepant results among the systems have been reported^9,10^. In addition, access to these automated systems and platforms can be limited for new antibiotics and settings^11^ with low laboratory resources such as research labs, outpatient clinics, point of care, and middle- and low-income countries.

**Table 1.**
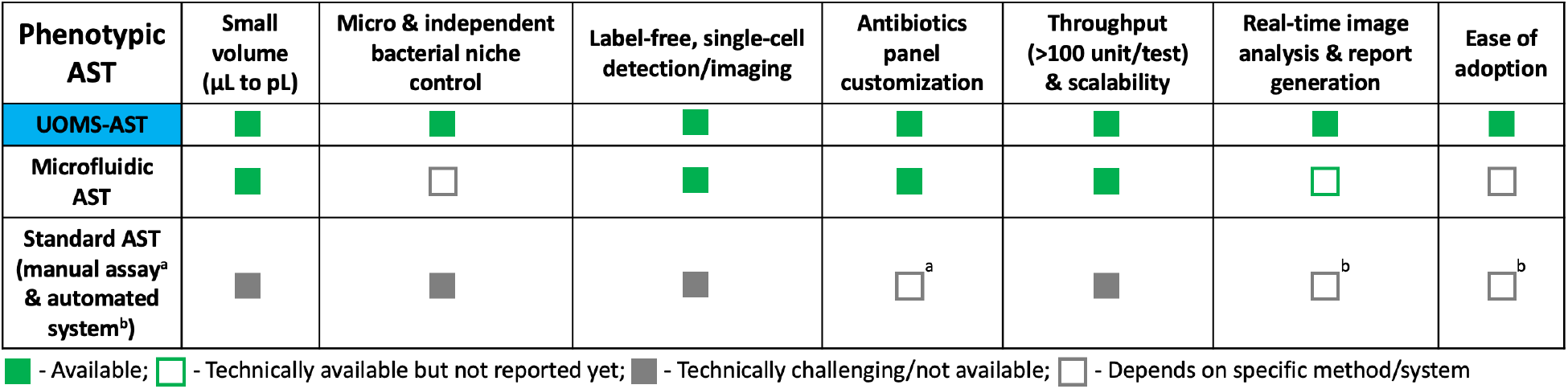
A comparison matrix showing the novel features of UOMS-AST compared to other phenotypic AST approaches/systems.

Recently, there has been renewed interest in the development of multi-liquid-phase microfluidics, named UOMS^12–25^. In UOMS, cell culture is implemented with the culture media and cells contained under an oil overlay, separating the cell culture/detection microenvironment from the ambient with an immiscible liquid (i.e., oil) rather than the closed chambers/channels of traditional microfluidic devices. Uniquely, UOMS allows: i) free physical access to the system with minimized evaporation and sample contamination, and ii) high-resolution optical access with various microscopic techniques (e.g., bright-field^13–16,18^, epifluorescence^14,16,20^, confocal^22^, multi-photon^21^). When combined with improved data analytics and reporting, UOMS has promising capabilities as a next-generation AST platform with significant advantages over standard and microfluidic AST methods (Table 1).

In this work, we demonstrate the ability of UOMS-AST to accurately and sensitively detect antimicrobial activity. Further, we demonstrate the integration of UOMS with cloud lab techniques for real-time image analysis and report generation, which allows automated, rapid phenotypic AST with application toward standard clinical laboratory settings.

## Results

### The UOMS platform

Since the 1980s, microfluidics and micro total analysis systems have rapidly grown and are now used in many commercial products^26,27^. Compared to closed-chamber/channel microfluidic systems, UOMS (Fig. 1a) utilizes surface chemistry contrast [i.e., free of physical structures (Fig. 1b, Supplementary Fig. 1)] and an oil barrier to confine aqueous media and cellular/molecular samples^16,20^ (Methods).

**Fig. 1.**
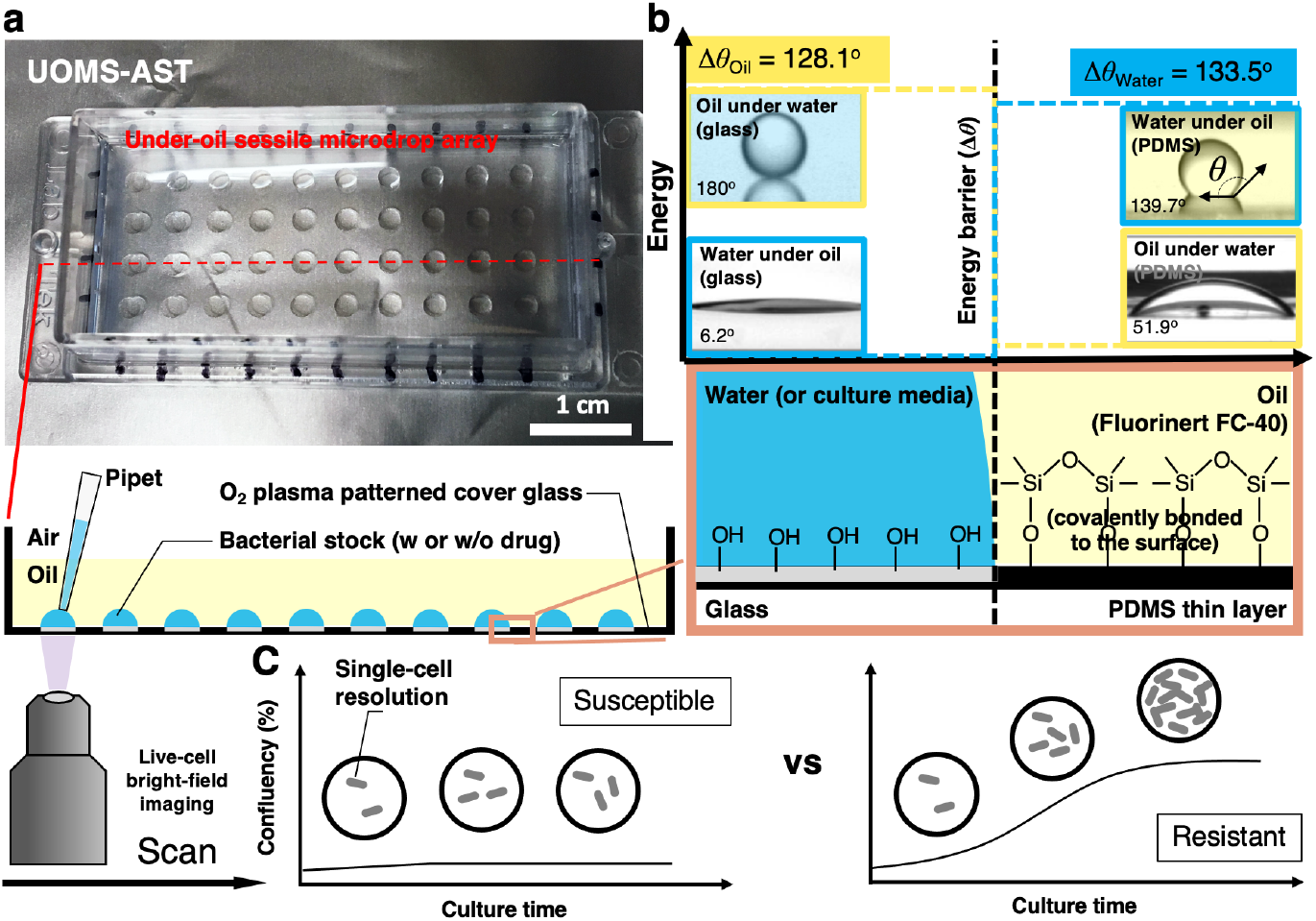
The configuration and physics of UOMS-AST. **a**, A UOMS-AST device fabricated with a 1-well chambered coverglass (6 cm × 2.4 cm, 0.16-0.19 mm in thickness). The glass surface was patterned for an array (4 × 10) of under-oil (fluorinert FC-40, or FC40) sessile (i.e., surface-attached) microdrops (2 mm in diameter). 2 μL of bacterial stock with or without antimicrobial was inoculated to each spot under oil by pipetting. **b**, A schematic shows the surface chemistry contrast for liquid confinement with the energy (or virtual) barrier (Supplementary Fig. 1). The glass surface was modified by a monolayer of covalently bonded, polydimethysiloxane (PDMS)-silane molecules and then selectively patterned by oxygen plasma. **c**, A schematic shows the live-cell, bright-field imaging in UOMS-AST and the detection of antimicrobial susceptibility and resistance based on confluency of bacterial cells in the field of view.

Specifically in the under-oil approach, we use fluorinated oil and/or silicone oil as the oil barrier, which minimizes oil extraction of lipophilic molecules [see the ultra-performance liquid chromatography-tandem mass spectrometer (UPLC-MS) characterization in our previous publication^22^]. The testing spots (i.e., plasma-treated areas) are hydrophilic [with an under-oil (fluorinated oil) water contact angle (CA) *θ* = 6.2°] and the untreated background hydrophobic (with an under-oil water CA *θ* = 139.7°). Similarly, the untreated background is oleophilic (with an under-water oil CA *θ* = 51.9°) and the plasma-treated areas oleophobic (with an under-water oil CA *θ* = 180°). To each liquid, the CA differential (Δ*θ*_water_ = 133.5°, Δ*θ*_oil_ = 128.1°) provides the energy barrier that holds and stabilizes the liquid on its preferred surface and thus enables a robust liquid-liquid boundary on the patterned, non-textured surface. Small volume (e.g., 2 μL in this work) of bacterial cells with or without antibiotic can be directly inoculated to the device under oil by regular pipetting. The coverglass bottom of the UOMS-AST device allows label-free (i.e., bright-field) optical access to the bacterial cells with single-cell resolution (Fig. 1c, Supplementary Fig. 2, Supplementary Movies 1-5). These configurations make the under-oil AST highly sensitive to pathogen’s antimicrobial activity.

### UOMS-AST

To validate the UOMS-AST method, we benchmarked it against standard phenotypic AST and antimicrobial assessments, i.e., broth microdilution-based growth and time-based killing assay (Methods). In this test, we used *Pseudomonas aeruginosa* (strain PA01) in a standardized inoculum (5 × 10^5^ cfu/mL) as a model human pathogen against four antibiotics with diverse mechanisms of action (Fig. 2a). The antibiotics were applied individually with different concentrations below and above their minimum inhibitory concentration (MIC). As shown, UOMS-AST was able to capture a full spectrum of different growth curves of the bacterium and displayed inhibition of growth at 0.5 µg/mL, consistent with the ciprofloxacin MIC in PA01 (Fig. 2b).

**Fig. 2.**
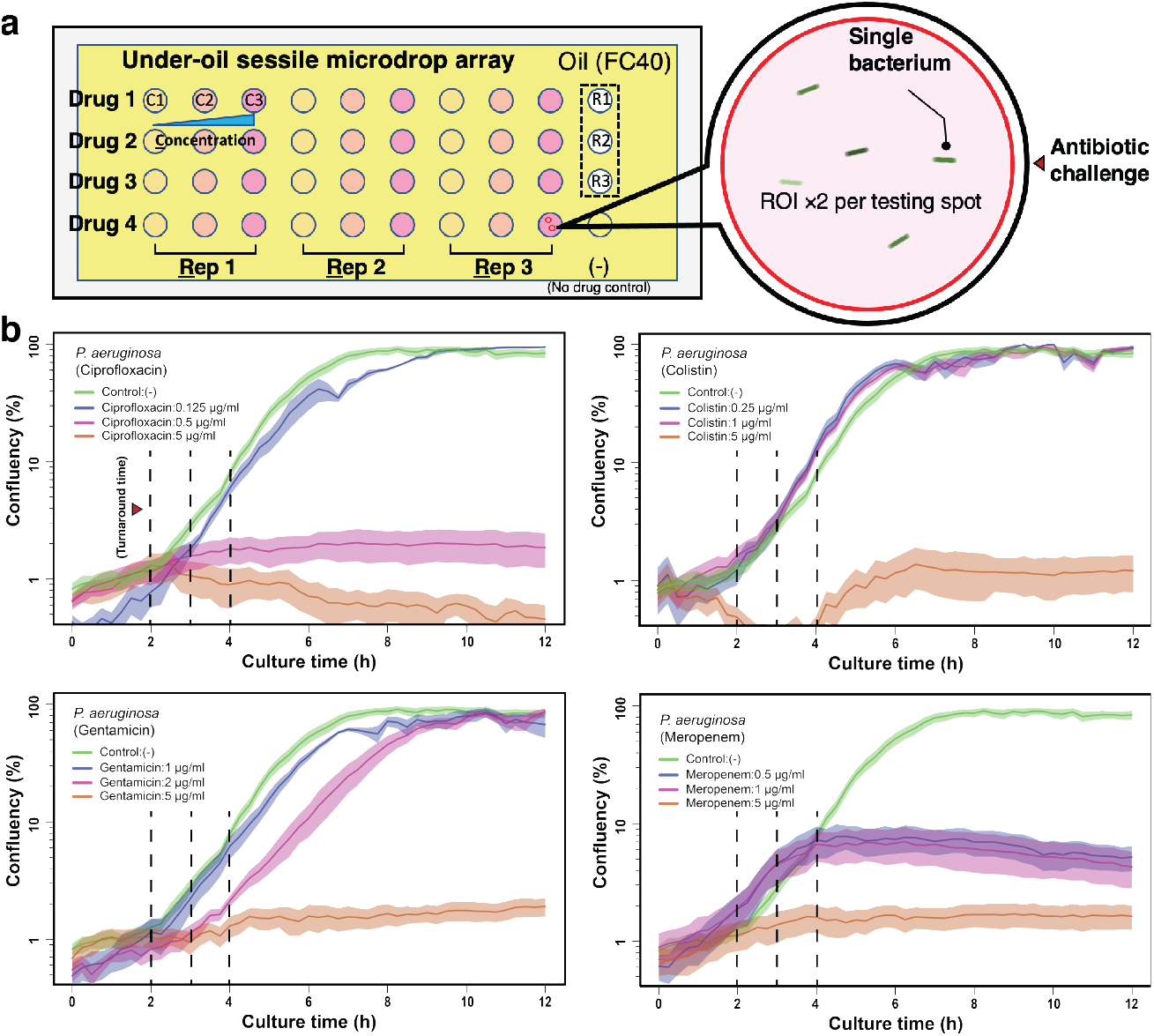
Results from UOMS-AST with *P. aeruginosa* (PA01). **a**, A schematic shows the layout of UOMS-AST on a chambered coverglass (6.2 cm × 2.4 cm) with 2 mm (in diameter) testing spots. Bacteria from 3 biological replicates (i.e., Rep 1 or R1, Rep 2 or R2, and Rep 3 or R3) were inoculated on the testing spots against four antibiotics (i.e., Drug 1, Drug 2, Drug 3, and Drug 4) at three different concentrations (i.e., C1, C2, and C3) and imaged under bright-field [60× magnification, ROI ×2 (the red circles) per testing spot] in a time lapse. **b**, The growth curves (i.e., confluency versus time, see Methods) of *P. aeruginosa* PA01 against four antibiotics. The antimicrobial activity and the difference across the conditions can be detected with a turnaround time of 2 to 4 h (vertical dashed lines). The solid line shows the mean of the biological replicates (×3) with the ROIs (×2) and the standard deviation (s.d.) is represented by the envelope on the plots. The microscopic images of bacteria are shown in Supplementary Fig. 2.

To compare the results from UOMS-AST against the Clinical and Laboratory Standards Institute (CLSI) approved standard, we performed broth microdilution susceptibility testing with strain PA01 against the same four antibiotics. PA01 MICs were ciprofloxacin 0.25-0.5 μg/mL, gentamicin 2 μg/mL, colistin 1-2 μg/mL, and meropenem 0.5 μg/mL. In comparison with the confluency results of the UOMS-AST (Fig. 2b, Supplementary Fig. 2), the antibiotic MIC susceptibility, determined at 16-20 h post incubation, aligned with the UOMS-AST data within the 2-4 h window (Fig. 2b, vertical dashed lines). To further compare UOMS-AST to the killing activity of antibiotics, we performed time-kill curve analysis as a standard assay for determining antimicrobial dynamic killing over time. Using the same organism and antibiotic sets, the time-kill assay (Fig. 3) replicates the UOMS-AST results for ciprofloxacin activity across the concentrations tested (0.125-5 µg/mL). Similar comparability was noted with the other antibiotics tested: gentamicin, colistin, and meropenem. These results indicate that UOMS-AST has utility for not only MIC phenotypic testing, but also for bactericidal concentration determination of antibiotics. Overall, compared to the standard phenotypic AST and the time-kill assay, the response to different antibiotics and doses in UOMS-AST can be identified automatically with a sample-to-answer turnaround time of 2 to 4 h.

**Fig. 3.**
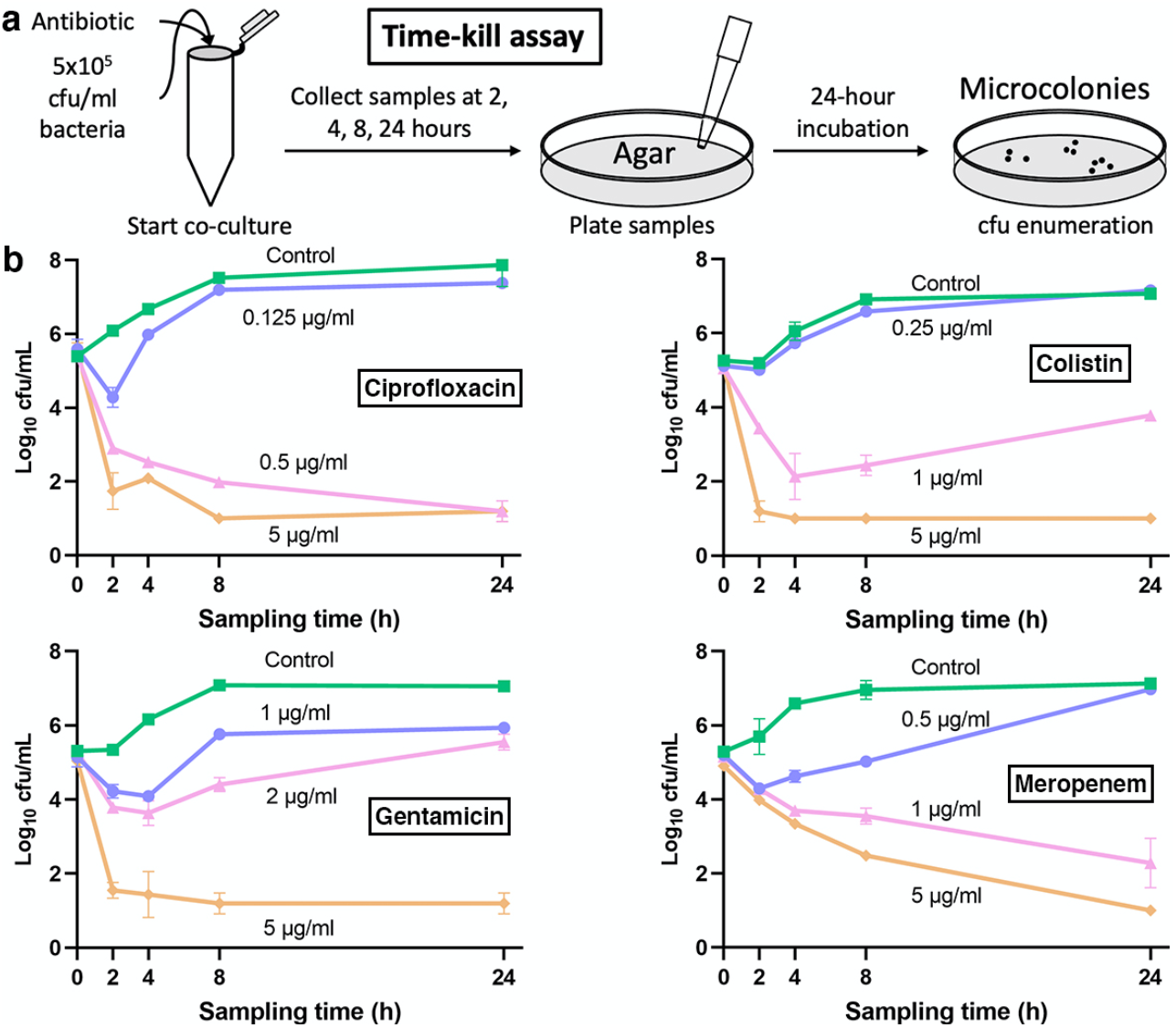
Standard time-killing curve results of *Pseudomonas aeruginosa* PA01. **a**, A schematic showing the time-kill assay workflow. In this assay, the bacteria were collected from each broth culture at a series of time points (i.e., the sampling times) and then streaked/cultured (up to 24 h) on an agar plate to obtain cfu count. **b**, The time-kill assay results of PA01 against the same antibiotics. Data were pooled and averaged with 3 replicates in each condition. Error bars, mean ± s.d.

### Cloud lab for real-time image analysis and report generation

Recently, various microfluidic platforms have been introduced to the field of phenotypic AST^28–31^. Distinct from the bulk-scale (i.e., agar or microtiter plate-based) phenotypic ASTs, microfluidic ASTs come with the capability of handling and processing ultra-small volume (microliter to picoliter) of samples, performing high throughput antimicrobial screening, and single-cell level antimicrobial detection sensitivity. However, it must be noted that often the reported turnaround time in microfluidics-based ASTs does not account for the time required for data analysis and report generation following data collection. Especially considering the data size (hundreds of GB to TB per run) from single-cell high throughput screening, the interpretation and quantification of the AST results can take days or weeks if done manually. An automated analytic and reporting mechanism in microfluidics-based ASTs that can be implemented with clinical laboratory standards is required to achieve clinically relevant, rapid sample-to-report turnaround times.

Here we demonstrate the combined use of a cloud lab-based, live cell imaging system in UOMS-AST (Fig. 4a, Supplementary Fig. 3, Methods). The system can be used in a standard cell culture incubator with controlled atmosphere (e.g., O_2_, CO_2_) and relative humidity. The UOMS-AST device in the workspace (i.e., a scanning area for the size of a standard microtiter plate) was scanned through a time course (Fig. 4b). A router transferred the images to cloud space during the scan. The image information in the defined region of interest (ROI) was analyzed with a selected algorithm, e.g., confluency in this work, and the report was generated in real time. Lab and healthcare personnel would have access to the recorded images and plots via the client installed on a personal computer or mobile device (e.g., smart phone).

**Fig. 4.**
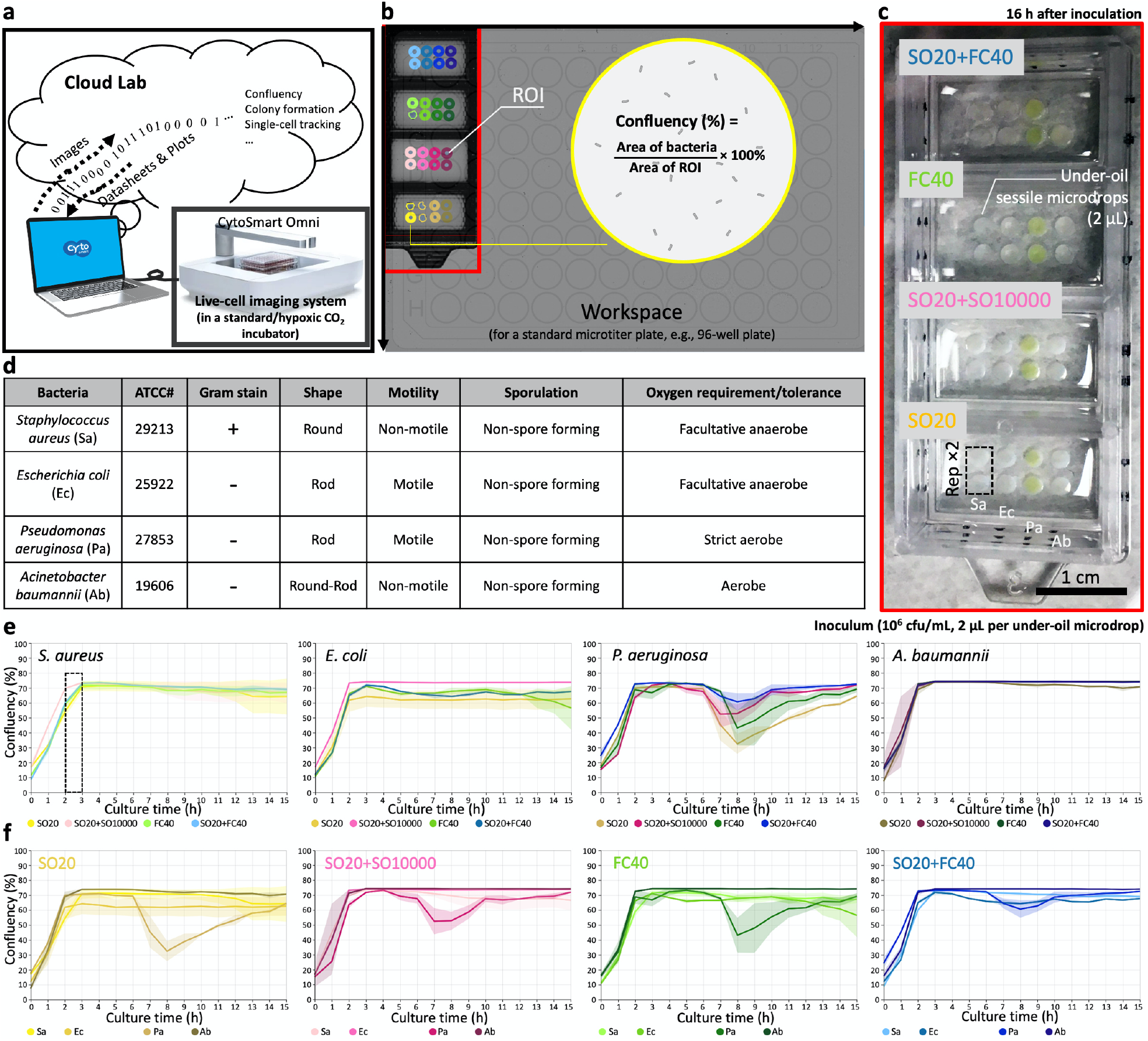
UOMS-AST integrated with cloud lab for real-time image analysis and report generation. **a**, A schematic shows the data flow of cloud lab. **b**, The workspace on CytoSmart Omni (Supplementary Fig. 3) can accommodate up to 6 pieces of (6 cm × 2.4 cm) chambered coverglass. Confluency of bacterial cells is automatically analyzed with the defined ROIs. **c**, A camera picture of a 4-well chambered coverglass device with bacteria and different oil overlays. Each condition (i.e., per bacteria species per oil condition) has two replicates (i.e., Rep ×2). **d**, A table shows the information of the four tested bacterial species. **e**, The growth curves (i.e., confluency versus time) of each bacterial species against the four different oil conditions. **f**, The growth curves of the bacteria species in each oil condition. The solid line shows the mean of the replicates and the s.d. is represented by the envelope on the plots.

We tested the capability of capturing the growth curve of four human pathogens - including *Staphylococcus aureus, Escherichia coli, Pseudomonas aeruginosa*, and *Acinetobacter baumannii* - that cause bacteremia and against four oil overlay conditions - including silicone oil (with the viscosity of 20 cSt, i.e., SO20), SO20+SO10000, FC40, and SO20+FC40 (Fig. 4c,d). We added silicone oil to this test because it provides three unique functions in UOMS: i) exclusive liquid repellency (ELR), where a liquid (e.g., culture media) is inherently and completely repelled by a solid surface (i.e., *θ* = 180°) when exposed to a secondary, immiscible liquid (e.g., oil) (see details in our previous publications^14,16,20^), ii) under-oil sweep distribution, where thousands of microdrops with a volume ranging from microliter to picoliter can be arrayed using automated or manual pipetting in a minute by dragging (or the so-called sweeping) a hanging drop of culture media (+ cells and/or drugs) across a patterned surface with double-ELR (i.e., under-oil water ELR + under-water oil ELR) (see details in our previous publications^16,20^), and iii) autonomously regulated oxygen microenvironments (AROM), where cells spontaneously set up, regulate, and respond to the oxygen kinetics via a supply-demand balance as seen *in vivo* (see details in our previous publication^22^). The double-oil conditions (i.e., one oil plus another oil) show the flexibility of adjusting the oil overlay by combining the properties of two oil types, e.g., different diffusion coeficcients of vital gases (e.g., O_2_ and/or CO_2_)^22^, or under-oil media evaporation/loss rate^14^. Specifically, SO20 allows smooth and robust under-oil sweep distribution due to its low viscosity and SO10000 can significantly reduce under-oil media evaporation/loss rate due to its ultra-high viscosity^32^. In addition, no visible oil-oil interface is generated when two silicone oils are used together in a system. The combined use of SO20 and FC40 allows both reliable under-oil sweep distribution and minimized under-oil media evaporation/loss rate due the ultra-low diffusivity and solubility of water molecules in fluorinated oil compared to silicone oil^33^.

As shown in the cloud lab results (Fig. 4e), the growth of the tested bacterial species were successfully recorded and quantified with real-time image analysis and report generation. All the four bacterial species reached confluence under oil within 3 h with a standardized inoculum (1 × 10^6^ cfu/mL). The four oil overlay conditions showed high consistency on the growth curves against each bacterial species (Fig. 4f).

## Discussion

Patients with serious bacterial infections (e.g., bacteremia) are at significant risk of complications, including antibiotic treatment failure^34^. Many studies have demonstrated that the most critical intervention to improve outcomes in patients with a severe infectious disease is early pathogen identification along with initiation of timely and effective antimicrobial therapy^35,36^. This “window of opportunity” is optimized in the first 24 h, e.g., in septic patients, and within 48-72 h in non-septic (but still severe) bacteremia patients. Importantly, studies have found that patients who are switched to appropriate therapy after receiving inappropriate initial therapy for bacteremia and sepsis are still at higher risk of poor outcomes^36,37^. Thus, rapid and accurate diagnosis is critical for all patients. Additionally, improved AST is a public health imperative to reduce unnecessary antibiotic use and hinder the risk of emerging antibiotic resistance.

Phenotypic ASTs provide functional readouts of antimicrobial activity and thus direct clinical guidance. Efforts have been seen in the development of next-generation phenotypic ASTs, striving to meet the following criteria: i) direct AST using the original clinical isolates (e.g., from blood, sputum, urine, abscess) to eliminate expansion culture that is time-consuming and introduces artificial passaging^8,38,39^, ii) rapid AST, i.e., fast sample-to-answer turnaround time, iii) comprehensive test coverage including anaerobes^40,41^, multispecies communities^42,43^, and novel phenotypes such as heteroresistance (i.e., resistant mutants within the wild type population)^44^, and iv) lower adoption/implementation barriers (e.g., alignment with clinical laboratory standards, small footprint, minimal personnel training, operation, and maintenance)^45^.

Microfluidics-based phenotypic ASTs have shown advantages over the traditional, bulk-scale methods in regard to detection sensitivity, speed, and throughput. Compared to the reported microfluidic platforms in this field, UOMS is naturally aligned with the standard tools (e.g., pipette, microtiter plate/chambered coverglass, inverted microscope) and lab automation (e.g., robotic liquid handler, 2D/3D cell printer) in biology and laboratory medicine^13–16,18,19^ where open systems are traditionally adopted. Due to the oil protection, small-volume (microliter to picoliter) ^16^ culture niches and/or testing units can be readily implemented with minimized evaporation and sample contamination. The small scale allows facile (i.e., only running xy scan without z stack), label-free (i.e., bright-field), live-cell imaging with single-cell resolution. Label-free, single-cell detection is critical in next-generation phenotypic ASTs because it minimizes the risk of introducing random artifacts/pre-selection from fluorescent transfection of the isolated pathogens, and maximizes the detection sensitivity and thus time efficiency. Importantly, the streamlined operation and small footprint of UOMS-AST make it suitable for translational applications in locations with limited space and access to infrastructures, e.g., mobile clinics and point-of-care settings.

Specifically, we highlight several unique features of UOMS-AST for its potential of commercialization. i) Low fabrication/material cost. The fabrication of UOMS devices is building upon room temperature (RT) chemical vapor deposition (CVD)^14^ and oxygen plasma surface patterning^16,20^ (Methods). The RT-CVD consumes minimal amounts of the surface-modifying reagents and energy. Both the surface modification and patterning are readily upscalable for mass production. Material-wise and compared to closed-chamber/channel microfluidic devices, UOMS completely removes the top and sides (e.g., plastic or elastomer) that are required in closed channel designs. Moreover, the PDMS stamps for oxygen plasma surface patterning^16,20^ are reusable and long lasting. Actually, the PDMS stamps in this work have been used for 2 to 4 years without showing any significant decay. ii) Easy storage and distribution. The UOMS-AST devices with a designated pattern can be stored in a sealed plastic foil package filled with a small volume of deionized water that covers the patterned surface. Based on the tests we have run in our lab, the shelf life with this storage method can be several months or longer. Before use, the device is dried with nitrogen gas and ready for use. iii) Low operating/environmental cost. Most microfluidic devices are designed or limited for one-time use. By contrast, the oil used in UOMS can be recycled, purified (e.g., by filtration), and reused. This will save the budget for the end users and in the meanwhile reduce the environmental burden of medical waste.

Our ongoing efforts in the further development of UOMS-AST are to fill the gaps related to the limitations of commercial AST methods. These include addressing the challenges in AST related to polymicrobial cultures, antimicrobial tolerance (including heteroresistance), inducible resistance, antimicrobial combinations, customization to new antibiotics, improving turnaround time (including direct from patient testing), and measuring bactericidal activity. Within these challenges, our first areas of focus are on direct AST, detection of heteroresistance with clinical isolates (i.e., pathogens directly isolated from clinical samples without expansion culture *in vitro*), and automated high-throughput (1000 to 10000 unit/test in nL to pL volume range of each testing unit) screening with both aerobes and anaerobes. The current gold standard for bacterial infection diagnosis and phenotypic AST consists of a complex, multi-step expansion culture process that can take days from sample to answer. Not only is it time consuming, but also expansion culture introduces artificial passage and selection to the original isolates making the detection of heteroresistance difficult (if not impossible). Recent studies have noted that heteroresistance is more prevalent among clinical infections than previously realized (up to 27%)(44), so having an assay to detect this phenotype is of critical importance to current patient care.

UOMS-AST, further combined with the three unique functions - ELR^14,16,20^, under-oil sweep distribution^16,20^, and AROM^22^ as described above in Results, provides a “less-is-more” strategy that leads to a next-generation phenotypic AST. The low adoption/implementation barriers give easy access to a significantly broadened group of end users, especially if commercialized. Importantly, the natural compatibility with clinical laboratory standards reserves the maximum lab resource efficiency and flexibility for different screening needs and tasks. Further, integrated with cloud lab techniques, automated, rapid, high-throughput antimicrobial detection and screening can be readily achieved.

## Methods

### Preparation of the UOMS devices

Detailed protocol was described in our previous publications^16,20^. Briefly, the process includes i) surface modification, ii) surface patterning, and iii) under-oil sample loading. Surface modition introduces a monolayer of covalently bonded PDMS-silane (1,3-dichlorotetramethylsiloxane, Gelest, SID3372.0) molecules by RT-CVD (Supplementary Fig. 1). The following surface patterning step is to transfer a designated pattern from a PDMS stamp (or mask) to the PDMS-silane grafted surface by selective oxygen plasma treatment. At last, oil is added to the device and cellular/molecular samples can be loaded under-oil by regular pipetting (the approach used in this work) or under-oil sweep distribution.

### Bacteria growth conditions and antibiotic susceptibility testing

Bacterial strains stored at -80°C were plated on Mueller-Hinton agar (MHA, BD Difco, BD, Franklin Lakes, NJ USA) and incubated at 37°C overnight prior to use. Time-kill assays were completed for *Pseudomonas aeruginosa* PA01 and the antibiotics colistin, meropenem, ciprofloxacin, and gentamicin (Sigma-Aldrich, St. Louis, MO, USA). The inoculum was prepared from a 0.5 McFarland standard and diluted 1:100 to obtain a 10^6^ CFU/mL starting bacterial concentration. The bacterial solution was aliquoted into microcentrifuge tubes and antibiotics were added at various concentrations. After 2, 4, 8, and 24 hours in a 37°C shaking incubator, a sample from each condition was collected, serially diluted, and spot-plated on MHA for CFU enumeration per ml. MICs were determined by broth microdilution, as described by CLSI^46^.

### Imaging and time lapse

We used Nikon Eclipse Ti (40× objective with 1.5× tube lens, i.e., 60× magnification) to acquire the bright-field images, and run the time lapse (15 min interval for 12 h) (Supplementary Fig. 2). The under-oil AST device was kept at 37 °C, 21% O2, 5% CO2, 95% relative humidity (RH) via an on-stage incubator (Bold Line, Okolab) during imaging.

### Batch-process image analysis and data visualization

We developed a custom image analysis workflow for objectively batch processing the time-lapse videos in JEX^47,48^, an open-source image analysis software that uses well-established libraries from ImageJ. Briefly, raw masks identifying bacterial cells were generated in two ways to robustly accommodate variation in bright-field imaging. In the first approach, bright-field images were gamma adjusted (γ = 0.7), inverted, background subtracted, Gaussian-mean filtered, and thresholded. In the second, images were background subtracted, inverted, Gaussian-mean filtered, and thresholded. The two masks were then combined using an OR operation to form the final mask. The surface area contributed by the bacterial objects were then extracted in JEX and then exported to.csv for analysis in R/RStudio. For data visualization, plots with confluency (%) over time were smoothed using the ‘smooth.spline’ function of the R ‘stats’ package with a smoothing parameter spar = 0.4.

### Cloud lab

The live-cell imaging system (CytoSmart Omni) adopted in the work is designed for use in a standard cell culture incubator with controlled temperature, atmosphere, and humidity (Supplementary Fig. 3). The system comes with 10× high-resolution camera mounted on a motorized xy actuator. The workspace is designed for a standard microtiter plate. It can take up to 6 pieces of the chambered coverglass used in this work at a time. The camera scans the workspace every hour. ROIs on the device can be set after the first scan or the whole time lapse scan. Image information from the ROIs will be analyzed by an algorithm (e.g., confluency) selected from the cloud lab. The user can have real-time access to the quantified results and plots with the client installed in a computer or mobile device.

### Cell line authentication

The bacterial cultures used in this study were purchased from ATCC. *Pseudomonas aeruginosa* Pa01 is a standard model organism for laboratory analysis, and strains *S. aureus* ATCC 29213, *E. coli* ATCC 25922, *P. aeruginosa* ATCC 27853, and *A. baumannii* 19606 are standard strains for CLSI antimicrobial susceptibility testing. For this study, these strains were authenticated for genus and species by selecting a single colony from overnight growth on solid agar and analyzing by MALDI-TOF MS according to clinical protocols and manufacturer’s instructions for identification (MALDI Biotyper, Bruker Corp., Billerica, MA). All five organisms were confirmed as correct.

### Statistical analysis

Raw data were directly used in statistical analysis with no data excluded. Data were present as mean ± s.d.. The replicate number was specified in the figure legends.

## Data availability

All study data are included in the article and/or supporting information. The data that support the findings of this study are available from the corresponding authors upon reasonable request.

## Code availability

The JEX workflows for batch-process image analysis and the R/R studio codes for data visualization are available upon request.

## Acknowledgements

This work was supported by NSF EFRI-1136903-EFRI-MKS, NIH R01 CA247479, NIH R01 AI154940, NIH R01 EB010039, NIH R01 CA185251, NIH R01 CA186134, NIH R01 CA181648, NIH R01AI132627, NIH P30CA014520, EPA H-MAP 83573701, and American Cancer Society IRG-15-213-51. We thank the following individuals for their assistance with this work: Dr. Taylor Fleet for sample collection, Dr. Brittney Hynes for bacterial cell line authentication, and Dr. Duane S. Juang for introducing the live-cell imaging system.

## Author contributions

C.L. conceived the method. C.L., and W.E.R. designed the research. C.L., and S.M. performed the experiments. C.L., J.W.W., S.M., C.F.V., and Z.H. performed the data analysis, interpretation, and visualization. C.L., D.R.A, W.E.R., and D.J.B. supervised the project. C.L. and W.E.R. wrote the manuscript and all authors revised it.

