## Supplementary Information for "Application of under-oil open microfluidic systems for rapid phenotypic antimicrobial susceptibility testing"

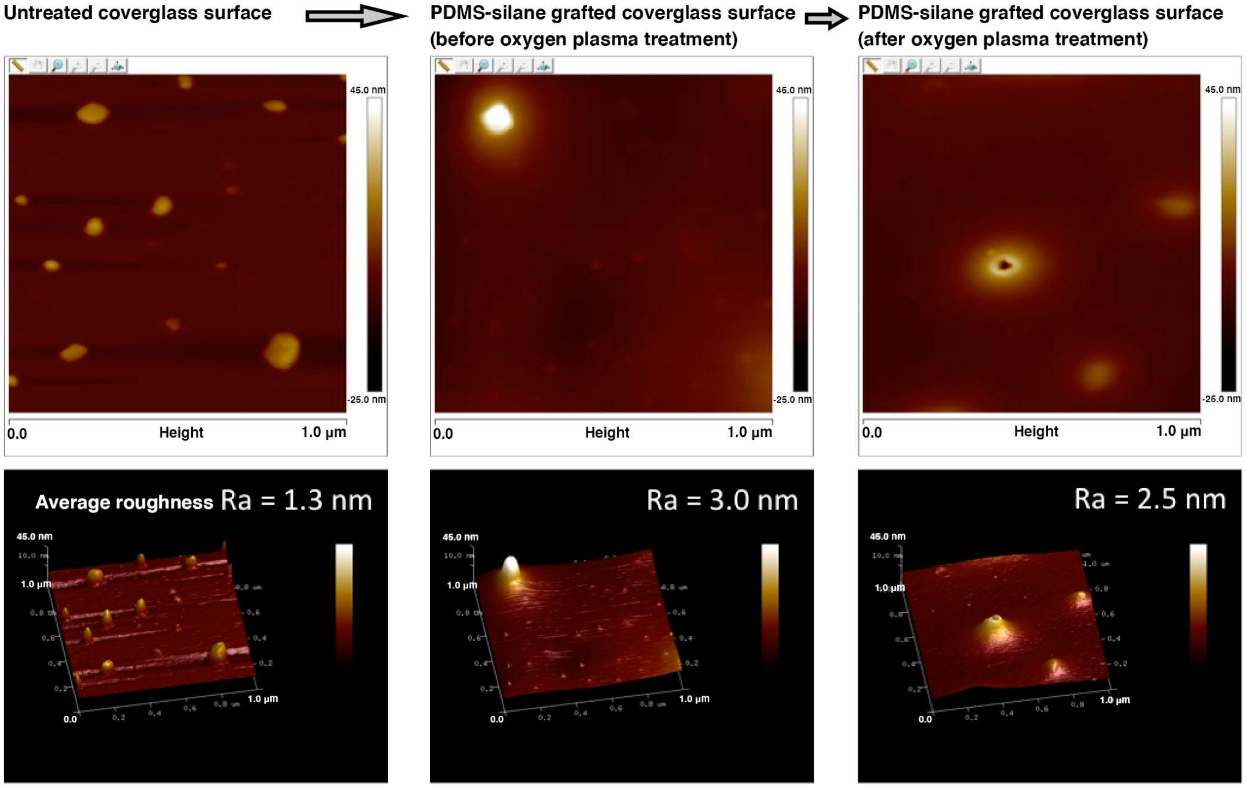


**Supplementary Fig. 1 Atomic force microscopy (AFM) characterization of a glass surface before and after PDMS-silane RT-CVD treatment and oxygen plasma treatment for surface patterning.**

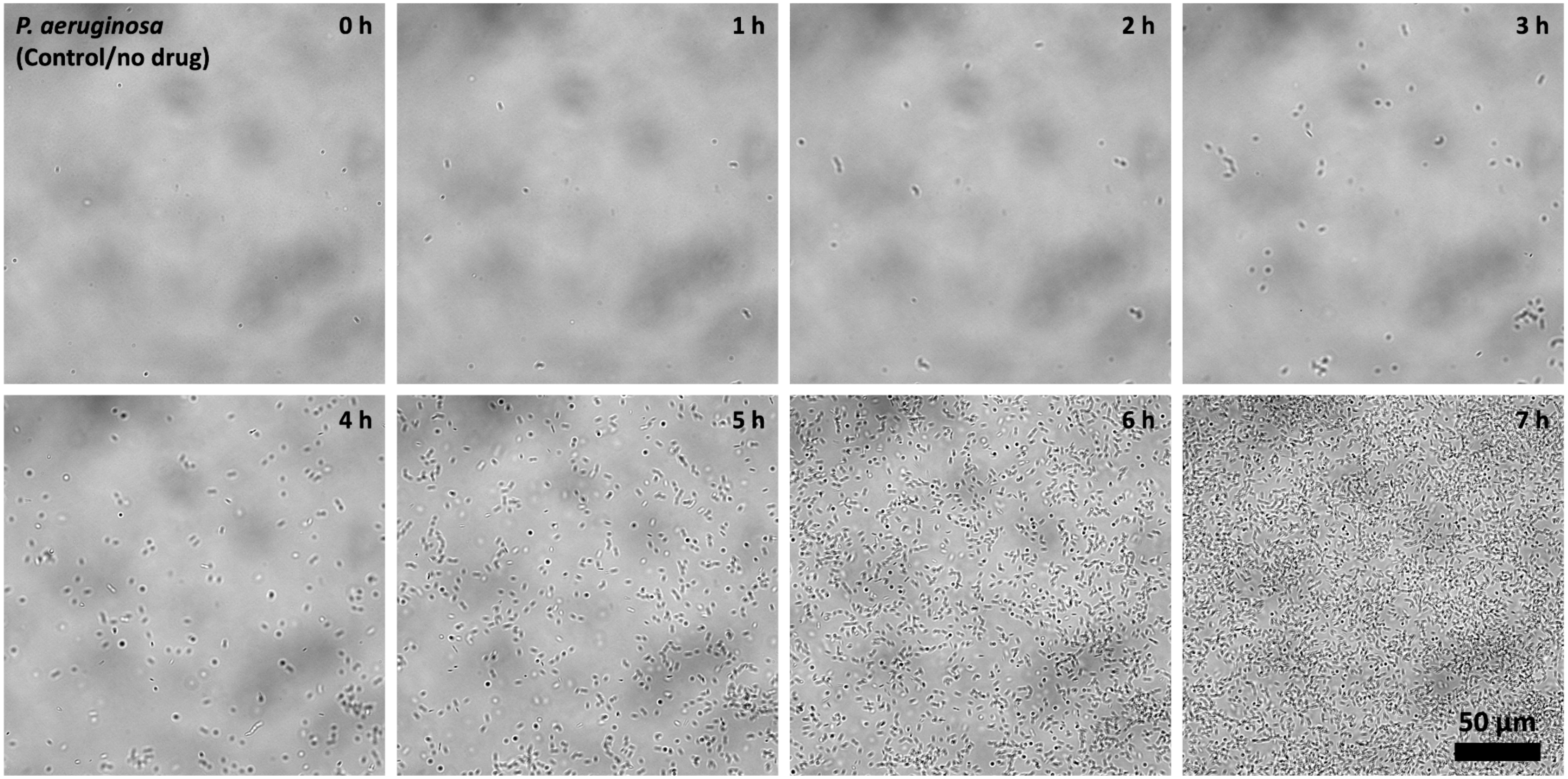
P. aeruginosa
(Control/no drug)
0 h
1 h
2 h
3 h
4 h
5 h
6 h
7 h
50 µm

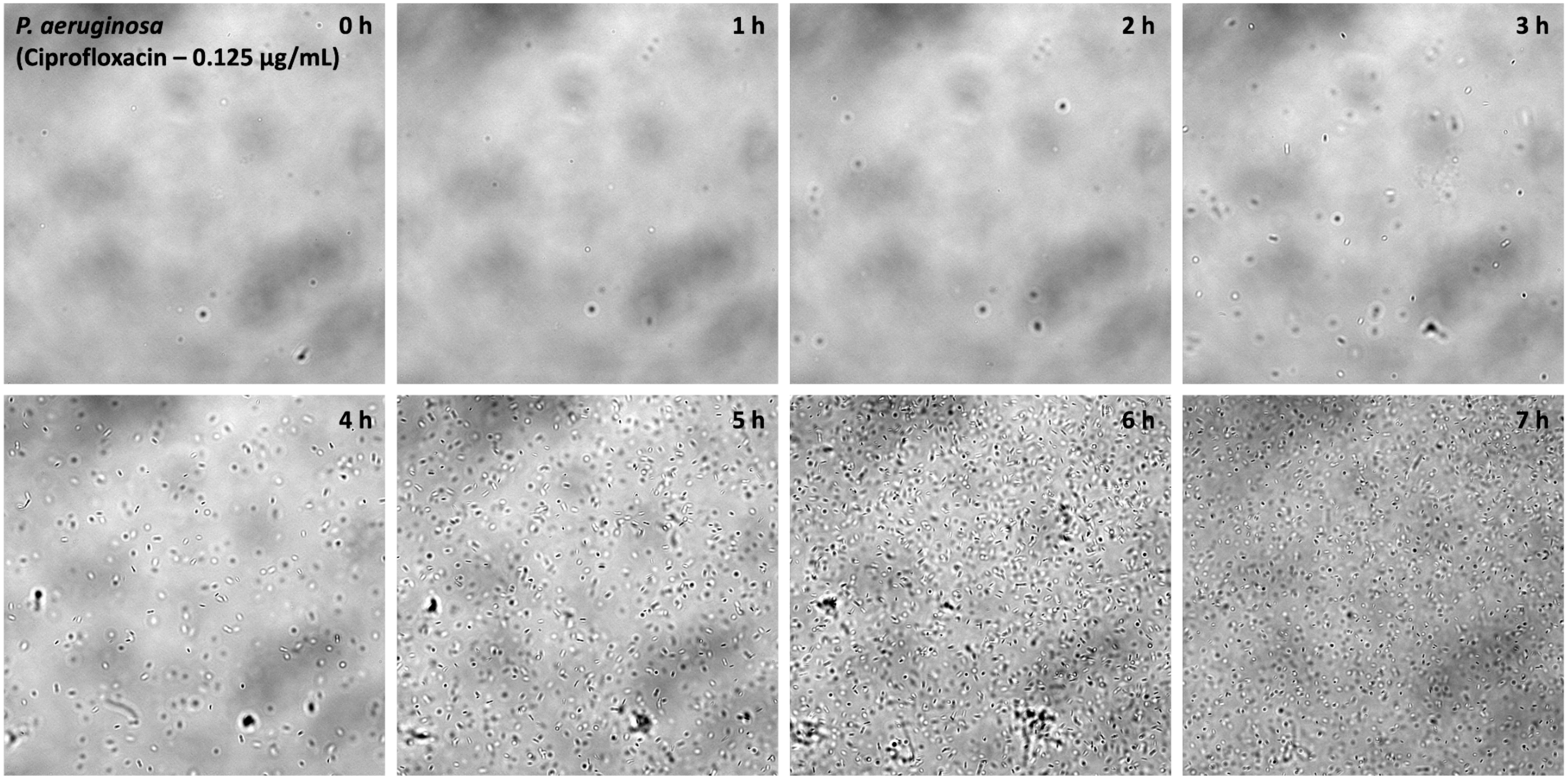
P. aeruginosa
(Ciprofloxacin – 0.125 µg/mL)
0 h
1 h
2 h
3 h
4 h
5 h
6 h
7 h

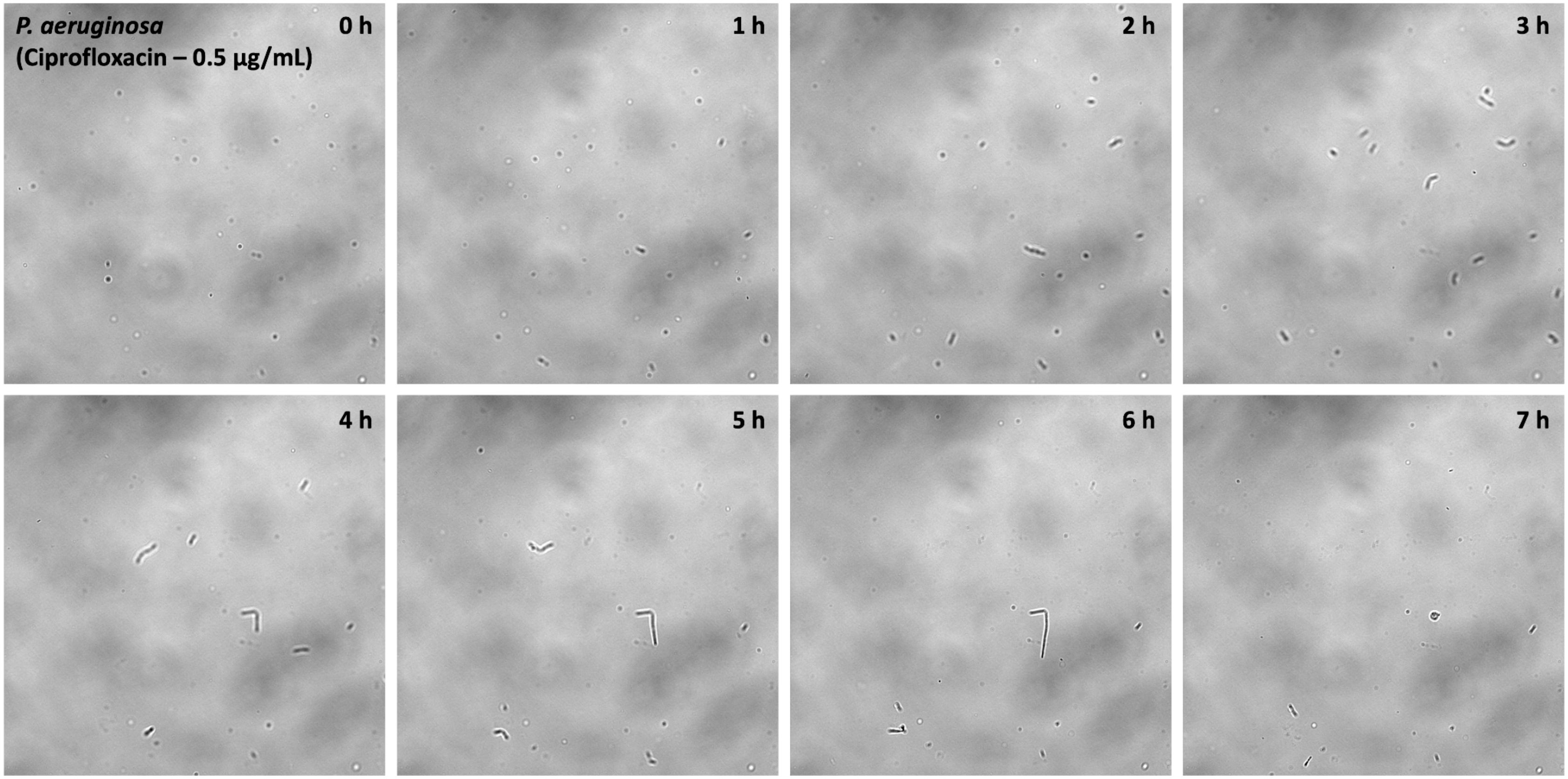

*P. aeruginosa*
(Ciprofloxacin – 0.5 µg/mL)
0 h
1 h
2 h
3 h
4 h
5 h
6 h
7 h

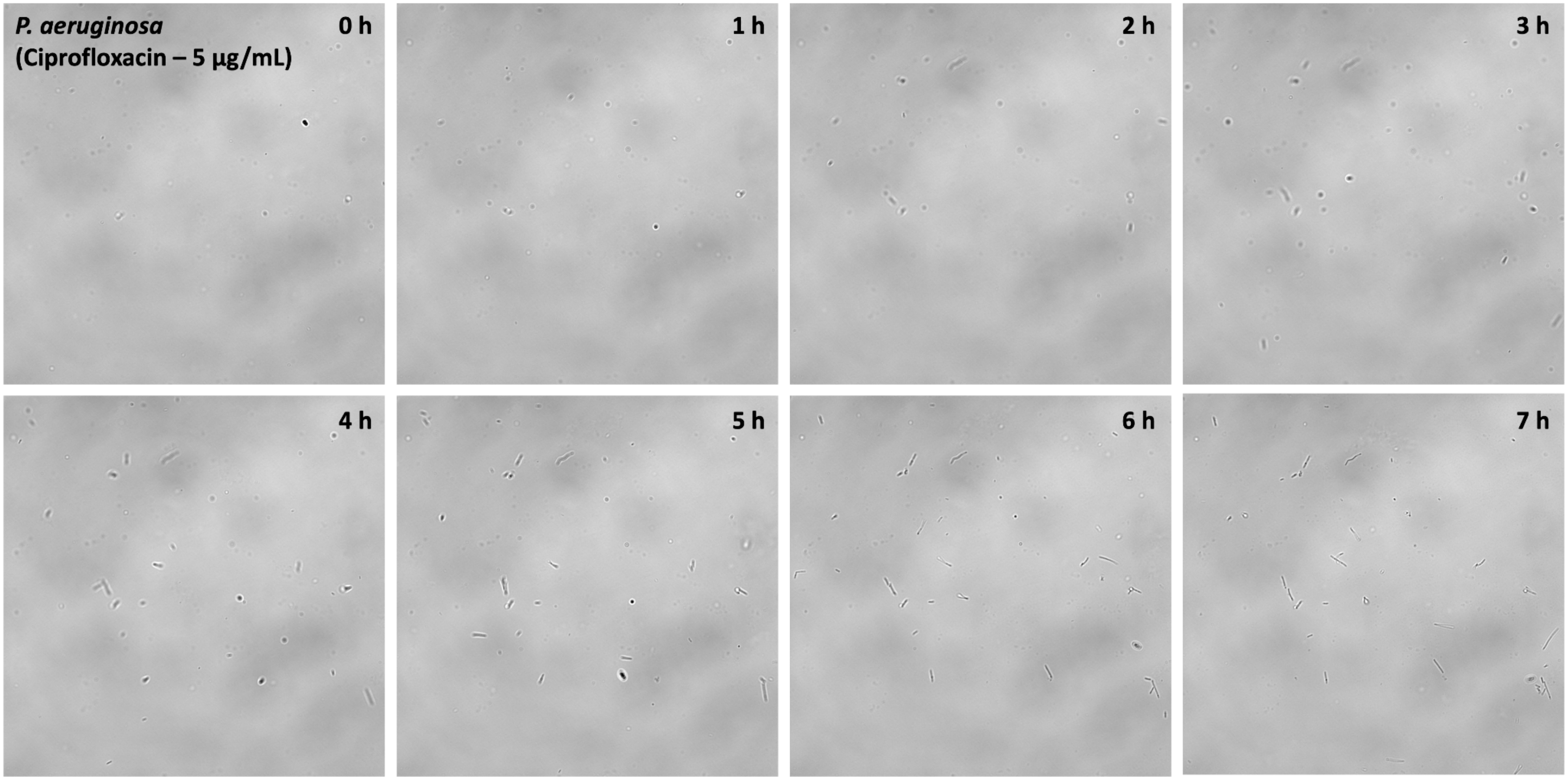

P. aeruginosa
(Ciprofloxacin – 5 μg/mL)
0 h
1 h
2 h
3 h
4 h
5 h
6 h
7 h

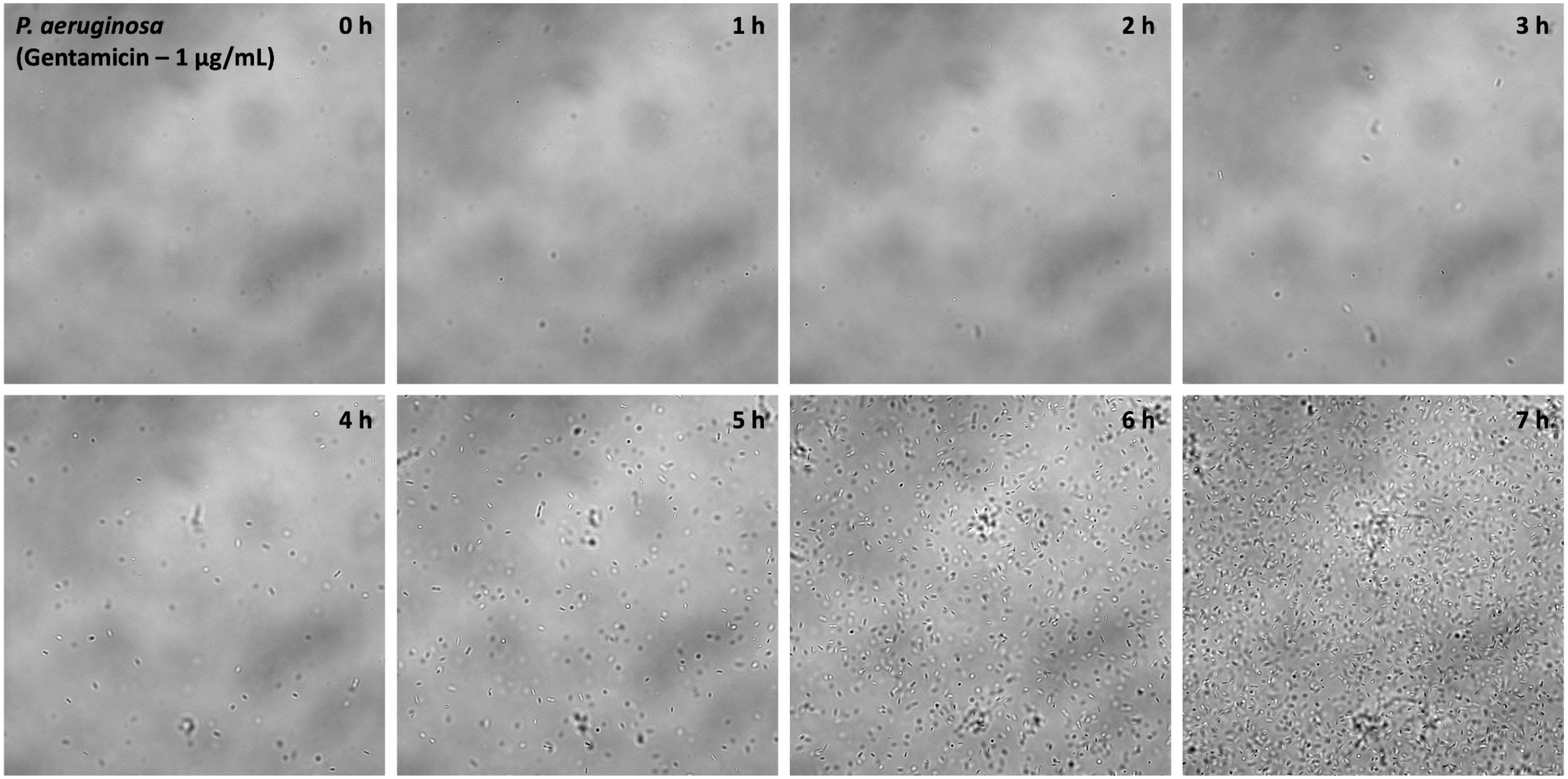
P. aeruginosa
(Gentamicin – 1 μg/mL)
0 h
1 h
2 h
3 h
4 h
5 h
6 h
7 h

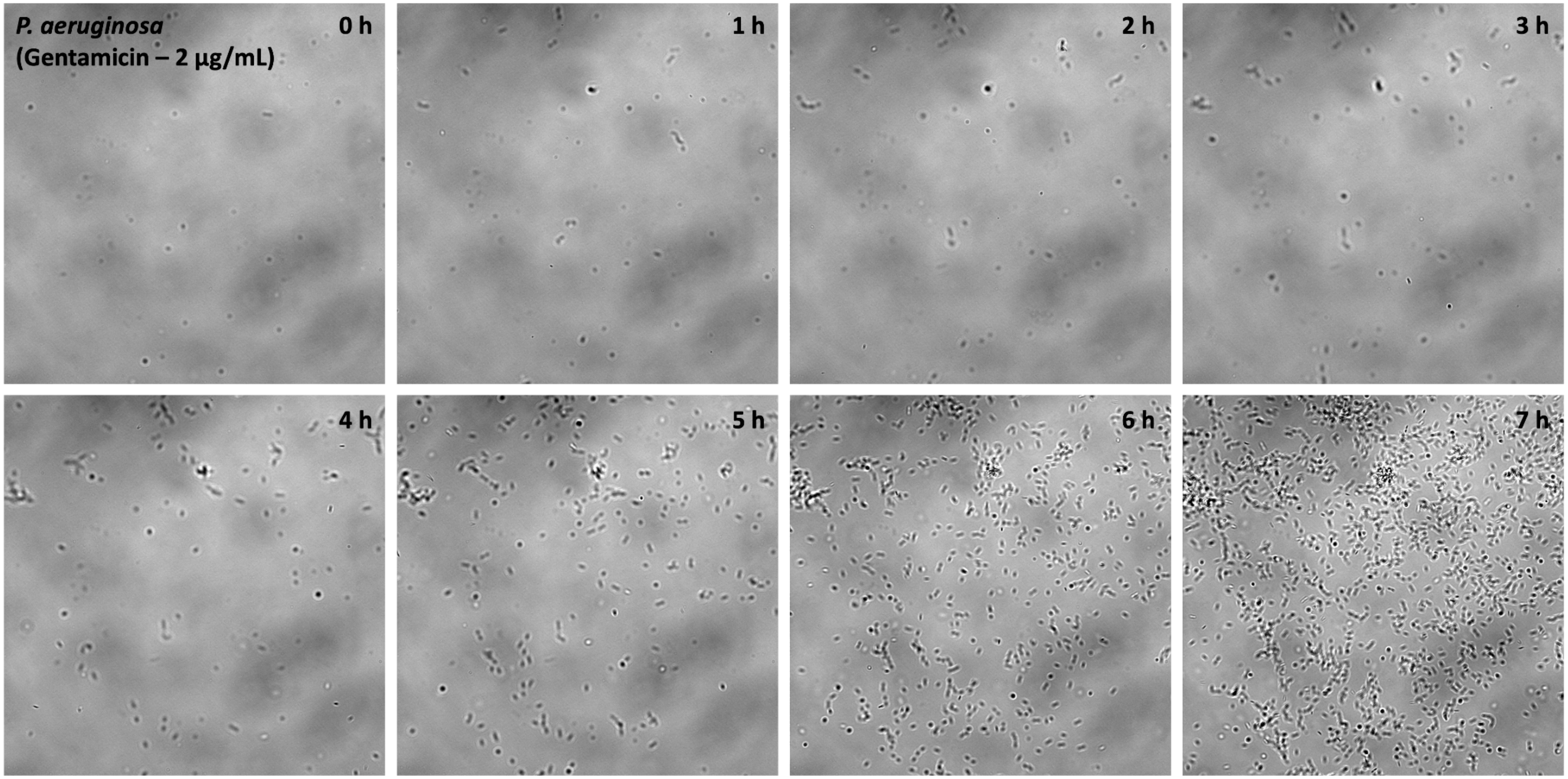
P. aeruginosa
(Gentamicin – 2 µg/mL)
0 h
1 h
2 h
3 h
4 h
5 h
6 h
7 h

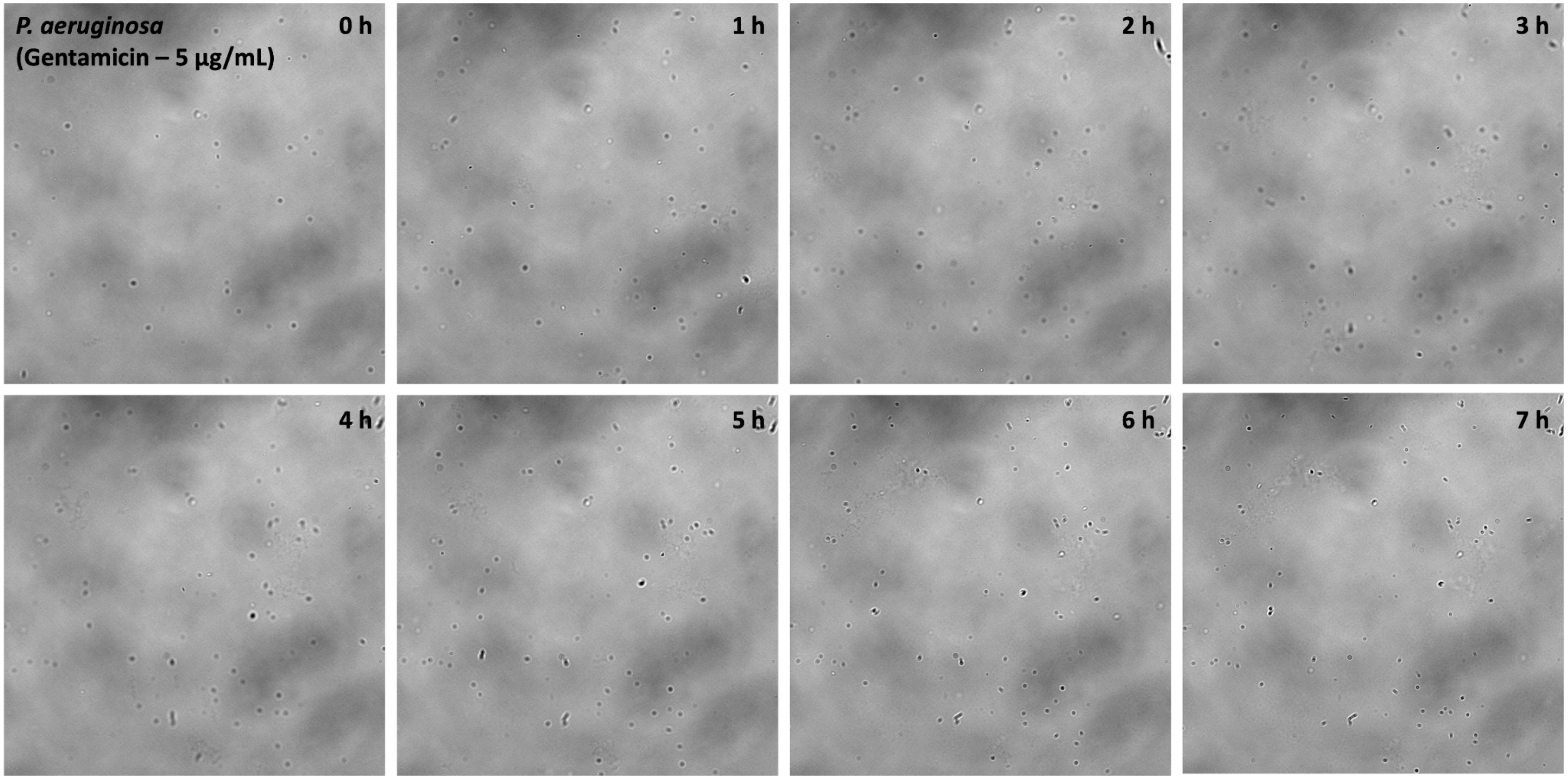

*P. aeruginosa*
(Gentamicin – 5 µg/mL)
0 h
1 h
2 h
3 h
4 h
5 h
6 h
7 h

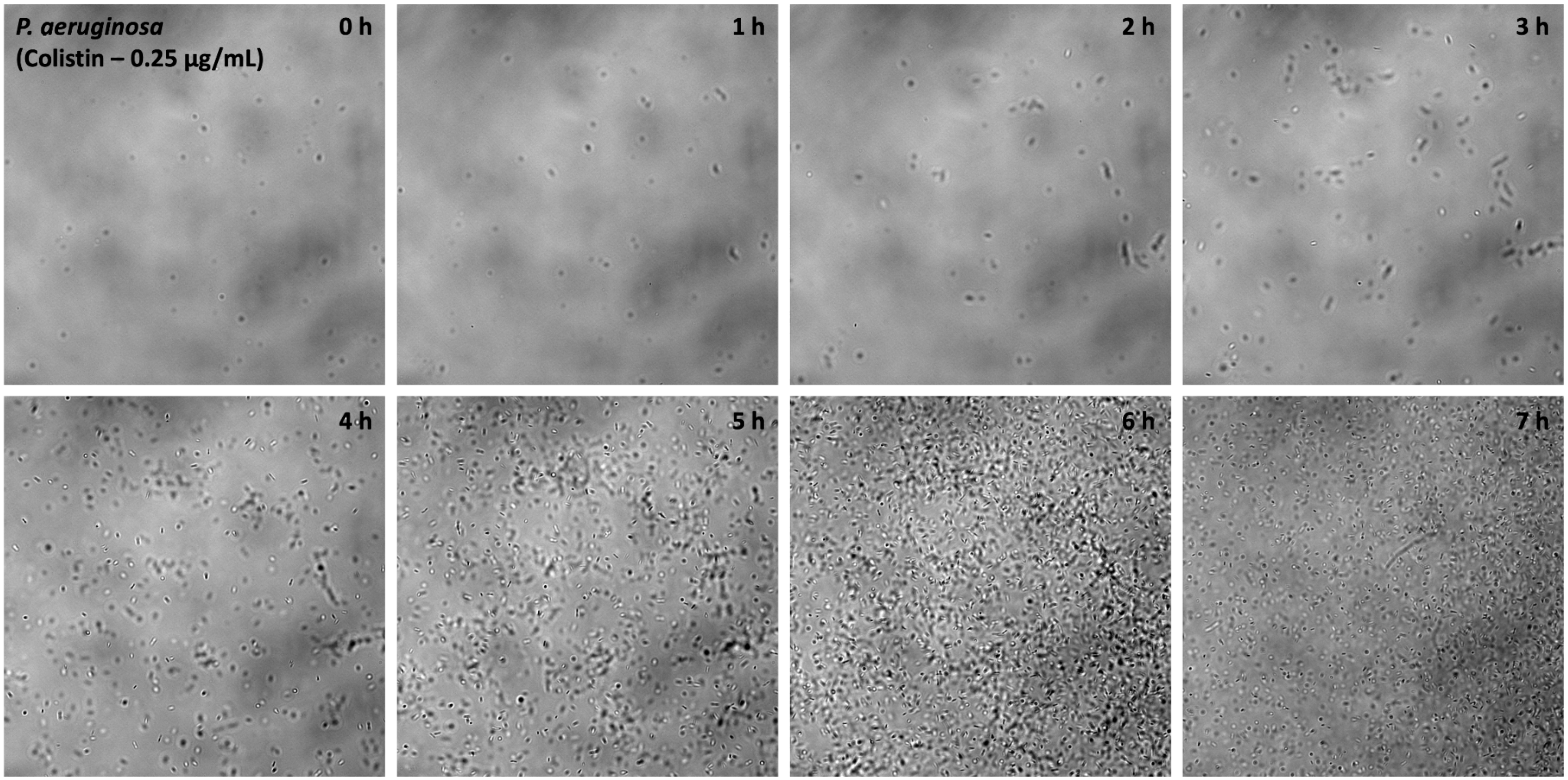
P. aeruginosa
(Colistin – 0.25 µg/mL)
0 h
1 h
2 h
3 h
4 h
5 h
6 h
7 h

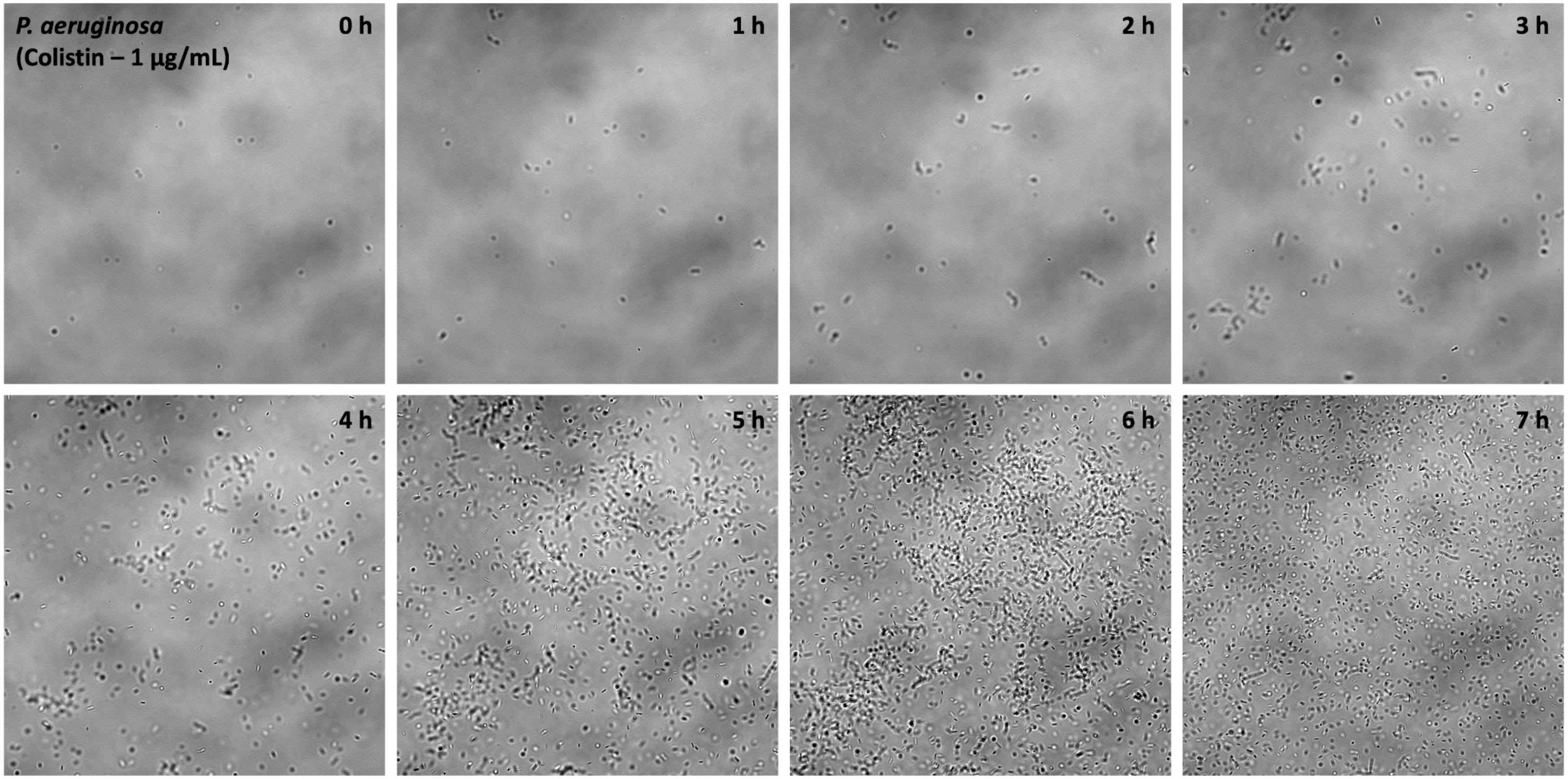
P. aeruginosa
(Colistin – 1 μg/mL)
0 h
1 h
2 h
3 h
4 h
5 h
6 h
7 h

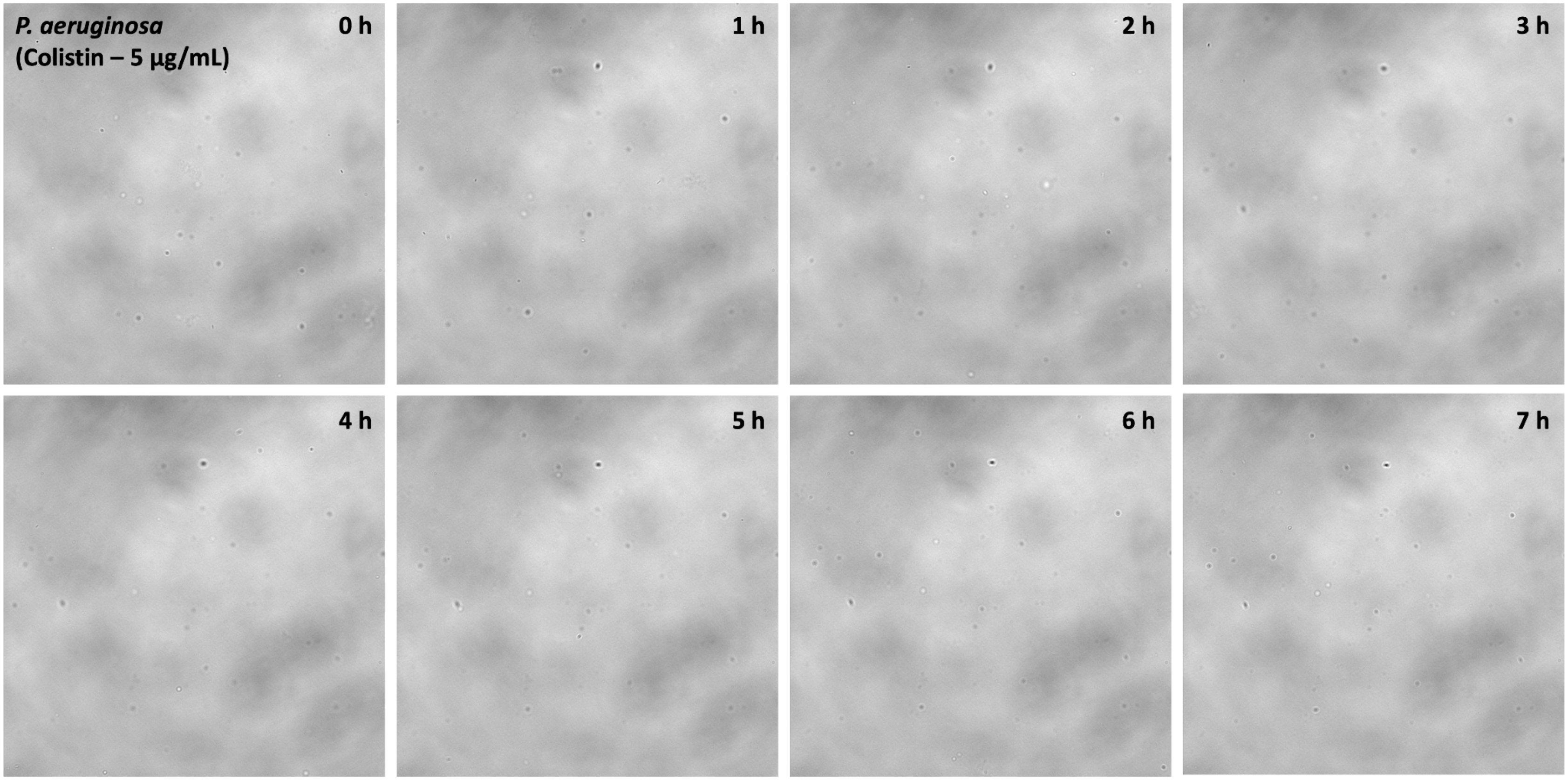

P. aeruginosa
(Colistin – 5 µg/mL)
0 h
1 h
2 h
3 h
4 h
5 h
6 h
7 h

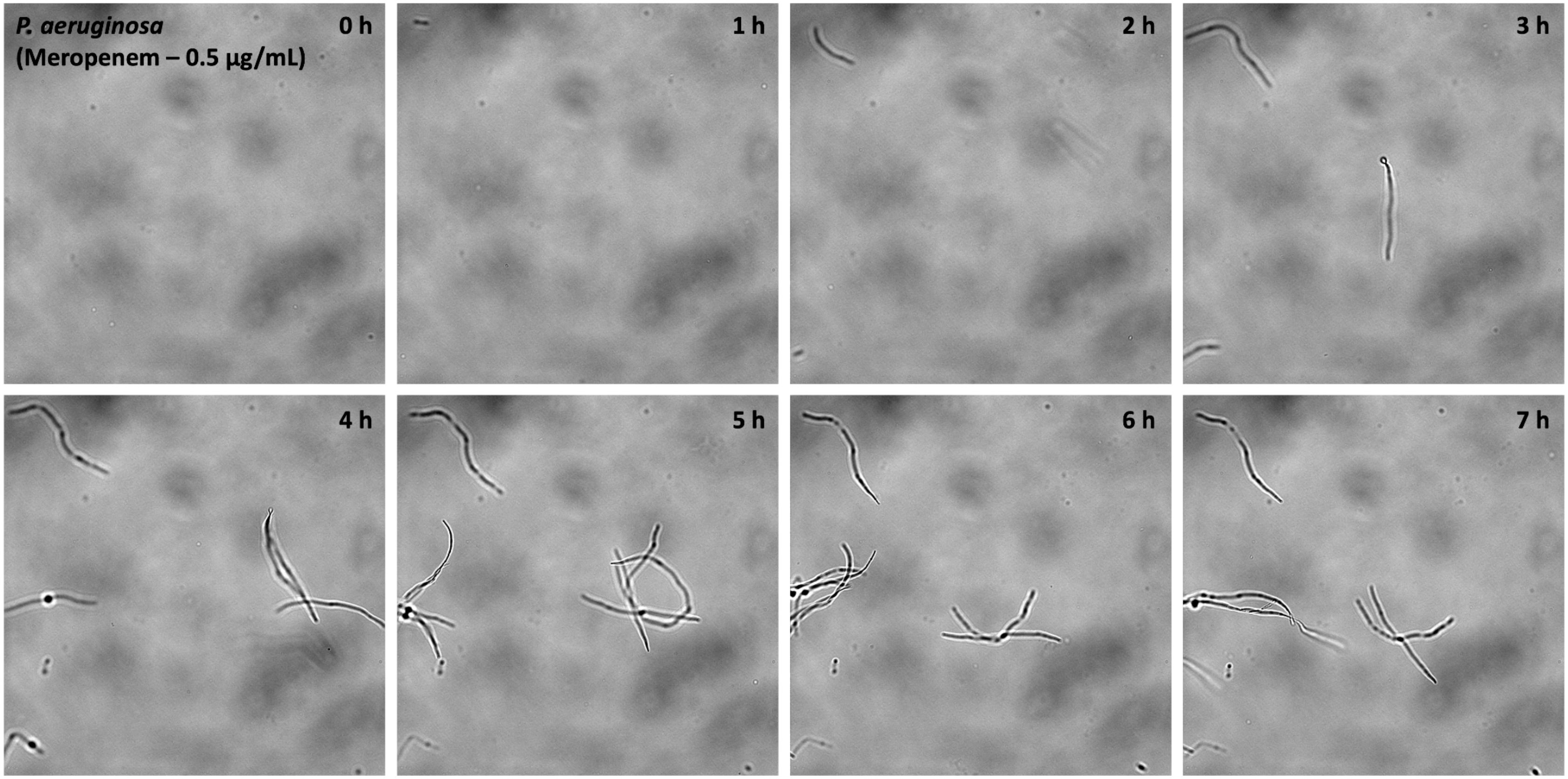
P. aeruginosa
(Meropenem – 0.5 µg/mL)
0 h
1 h
2 h
3 h
4 h
5 h
6 h
7 h

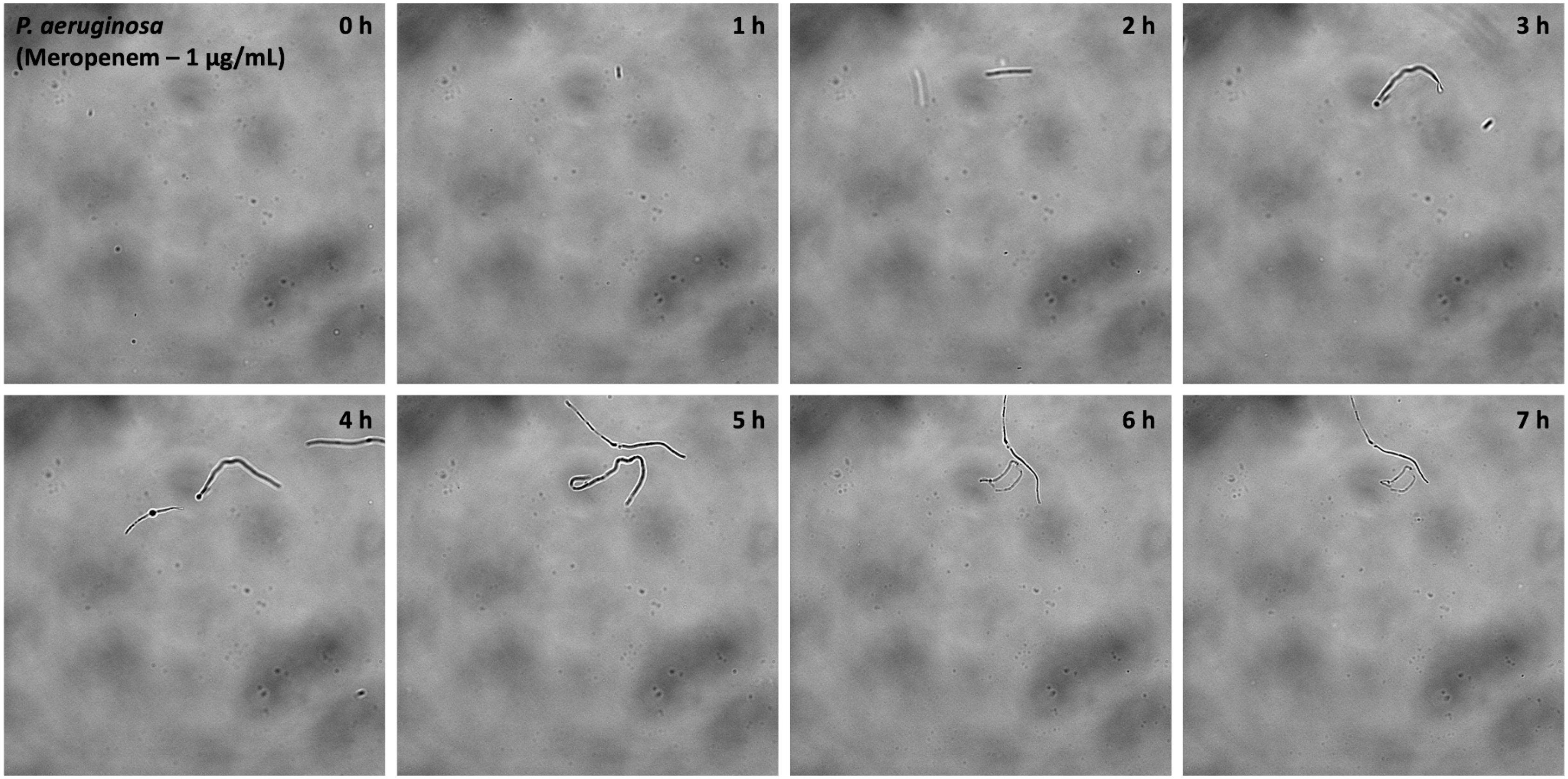
*P. aeruginosa*
(Meropenem – 1 µg/mL)
0 h
1 h
2 h
3 h
4 h
5 h
6 h
7 h

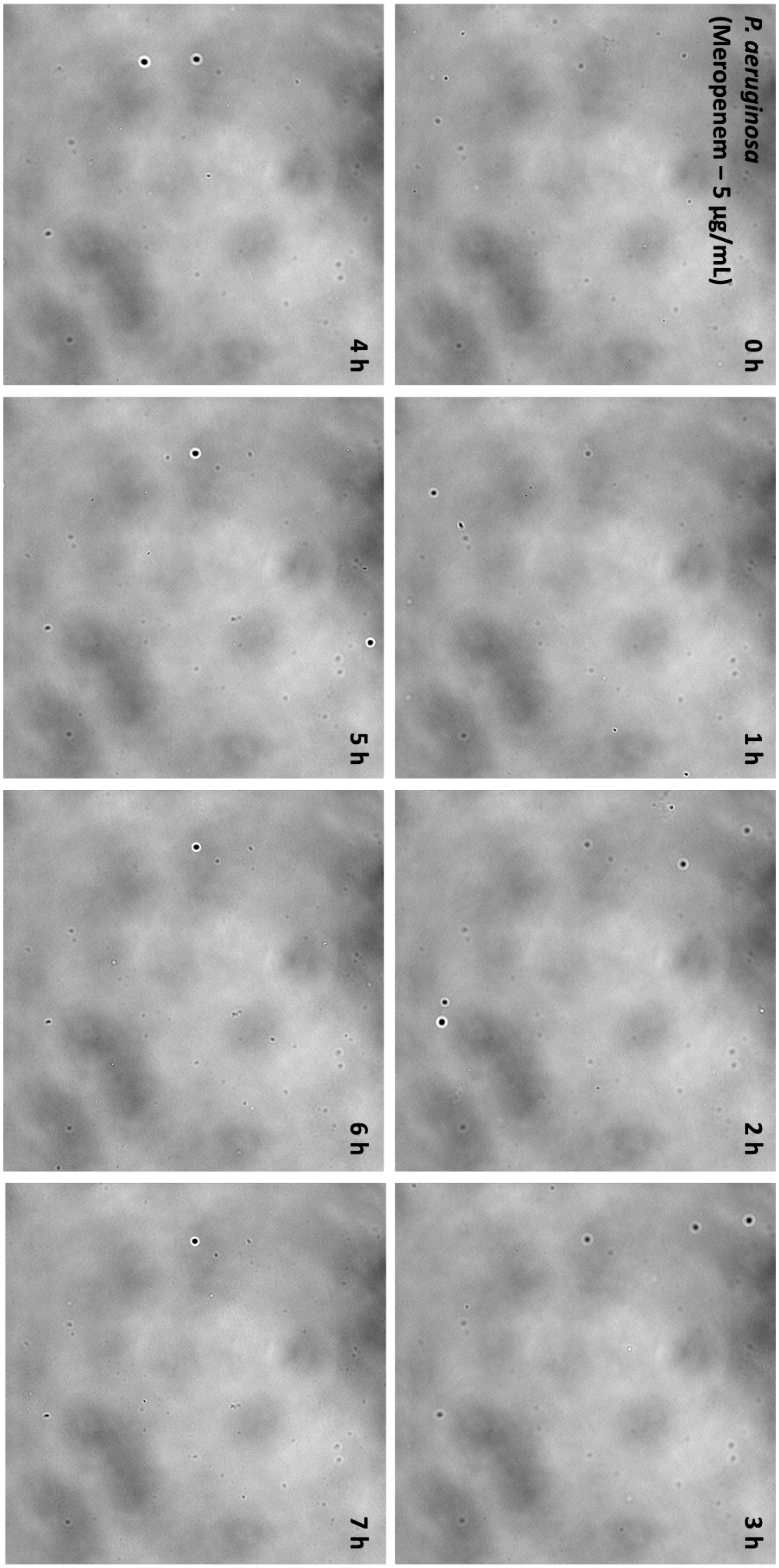


**Supplementary Fig. 2 Bright-field image (60× magnification) sequence from the time lapse in Fig. 2.**

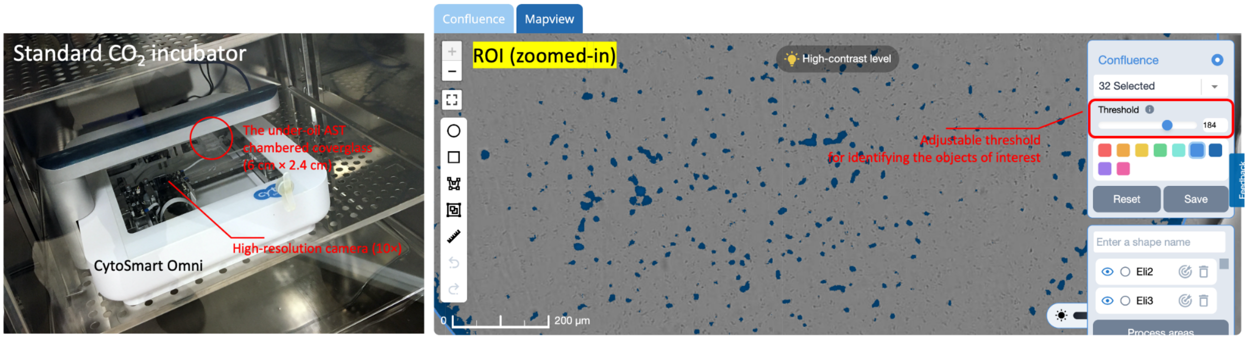


**Supplementary Fig. 3 The CytoSmart Omni system in a standard $CO_2$ incubator and the graphic user interface (GUI) for image thresholding and object identification in the confluency analysis.**

**Supplementary Movie 1 Time lapse (12 h, 60× magnification)_*P. aeruginosa* (PA 01)_Control no drug (3600× accelerated)**

**Supplementary Movie 2 Time lapse (12 h, 60× magnification)_*P. aeruginosa* (PA 01)_Ciprofloxacin 0.125 μg/mL (3600× accelerated)**

**Supplementary Movie 3 Time lapse (12 h, 60× magnification)_*P. aeruginosa* (PA 01)_Getamicin 1 μg/mL (3600× accelerated)**

**Supplementary Movie 4 Time lapse (12 h, 60× magnification)_*P. aeruginosa* (PA 01)_Colistin 0.25 μg/mL (3600× accelerated)**

**Supplementary Movie 5 Time lapse (12 h, 60× magnification)_*P. aeruginosa* (PA 01)_Meropenem 0.5 μg/mL (3600× accelerated)**
